# An online shared decision-making intervention for dementia prevention: A parallel group randomized pilot study

**DOI:** 10.1101/2023.07.16.23292730

**Authors:** Raymond L Ownby, Rosemary Davenport

**Author notes:** Corresponding author: Dr. Ownby, Center for Collaborative Research Suite 430, 3300 South University Drive, Fort Lauderdale FL 33314. Voice.

## Abstract

**OBJECTIVES:** The objective of this study was to evaluate the acceptability and efficacy of an online dementia prevention intervention based on a cognitive behavioral shared decision-making model.

**DESIGN:** This was an unblinded pilot study in which participants were randomly assigned to one of two treatment groups.

**SETTING:** This study was carried out remotely via telephone, video conferencing and online data collection.

**PARTICIPANTS:** Persons 50 years of age and older interested in developing more brain healthy lifestyles were recruited.

**INTERVENTION:** Both groups received 12 weekly sessions on lifestyle factors related to cognitive decline. The treatment as usual (TAU) group received the information and was encouraged to make lifestyle changes. The cognitive behavioral shared decision-making model (CBSDM) group received structured weekly sessions with support for evidence-informed personal goal choices and behavior change strategies.

**MEASUREMENTS:** Primary outcome measures were the Alzheimer Disease Risk Inventory and the Memory Self-Efficacy and Dementia Knowledge Assessment Scales. Participants reported brain health activities during the first, sixth, and 12^th^ weeks of the study.

**RESULTS:** The intervention was viewed positively by participants who all said they would participate in it again. Participants in the CBSDM group showed increases in knowledge of dementia risk factors and in exercise. Other outcomes were consistent with moderate to large effect sizes for both groups.

**CONCLUSIONS:** An online intervention providing psychoeducation and behavior change support was viewed positively by older adults. Results provides preliminary support for the CBSDM intervention’s efficacy in promoting brain health in older adults. (243 words)

**Registered at clinicaltrials.gov:** NCT04822129

## Introduction

Cognitive decline and dementing illnesses affect a large number of older persons, and that number is expected to increase worldwide over the next several decades. The clinical and economic impact of these conditions makes the search for effective treatments and preventive interventions critical. Although there have been breakthroughs recently in the treatment of Alzheimer’s disease (1), the need for interventions that can be broadly implemented continues to be significant. Several studies suggest, for example, that treatments that could prevent the onset of dementing illnesses or even delay their onset by several years would have significant economic benefits (2, 3).

A promising target for intervention has been a number of modifiable factors that could affect the risk for developing cognitive decline and dementia; one review concluded that intervening on modifiable risk factors could reduce the number of new cases of dementia by as much as 40% (4). Many of the most important risk factors are related to lifestyle, including exercise, diet, and cognitive activity. Even risk factors that are related to well-defined disease entities such as diabetes and hypertension have clear relations to lifestyle. Interventions that could affect these factors might be useful in reducing risks for cognitive decline and dementia in older adults. Further, several studies of multifactorial interventions such as the FINGER trial (5) have shown clear benefit in addressing multiple lifestyle factors.

While research suggests the potential benefits of lifestyle change for older adults, it is not clear that these benefits are clearly communicated to them. Popular works promising information on how to implement lifestyle changes in one’s own life vary in emphases on one or a small number of possible factors; few if any have been empirically tested (6). Large-scale surveys show that older adults are interested in brain health and willing to make changes to decrease their risk for cognitive decline and developing dementia (7, 8), but are not sure how to go about doing so (8-11). Even with this uncertainty, one study showed that even though half of participants believed they were at high risk for developing dementia, only 5.2% of them had discussed brain health with their physicians (12). Clearly, older persons can benefit from guidance on how to make lifestyle changes to support and improve their brain health. While the need for guidance is apparent, how to provide it is not.

One strategy for helping patients make health-related decisions with a substantial evidence base is shared decision making (13). Shared decision making occurs when a patient and provider work together to make a decision that incorporates the patient’s preferences and the provider’s expertise to arrive at a plan of action (13). Shared decision making has been related to greater patient satisfaction and better health outcomes (14). Elsewhere, we outlined a model for a dementia prevention intervention based on a shared decision model for brain health that integrates psychoeducation on risk and protective factors with coaching based in cognitive behavioral change techniques (15). The purpose of the current study was to evaluate the usefulness, acceptability, and efficacy of a model-based intervention deployed online with older adults.

## Methods

This was an unblinded randomized pilot clinical trial in which all participants completed baseline and follow-up evaluations and received a brain health intervention consisting of 12 weekly 30-40 minutes sessions, including an initial introductory and a final wrap-up and review session.

### Groups

The intervention delivered as treatment as usual (TAU) was created to be a reasonable representation of what many older persons face in finding out about the importance of brain health and making lifestyle choices if they choose to try to improve it. A great deal of information is available online and in popular books, and authors have proposed a number of programs for brain health (16-18) that recommend multiple activities or interventions with little guidance on prioritizing among them or strategies for behavior change. The TAU intervention mirrored this situation, providing participants with information resources from which to choose activities and methods of behavior change but without specific guidance on activity or behavior change strategies.

The intervention based on the cognitive behavioral shared decision making model, or CBDSM (15), included the same psychoeducational component as delivered to the TAU group, but the individual online sessions were structured to be similar to sessions of cognitive behavioral therapy (19), with an emphasis on collaborative goal setting, education in self-monitoring, and problem solving. The model-based intervention aimed to help older adults make evidence-informed choices about lifestyle factors that could help reduce their risk for cognitive decline and dementia. Grounded in the cognitive behavioral model of behavior change, it draws on a shared decision-making approach to working with older adults as healthcare providers act as consultants and facilitators of goal choice while taking individual patient needs, preferences, and circumstances into account. The model specifies three core elements for the intervention: (a) psychoeducation on risk and protective factors organized according to three hypothesized mechanisms by which they affect brain health, (b) additional education on evidence-informed approaches to goal choice, and (c) instruction in behavior change strategies with ongoing support.

The three mechanisms by which activities are hypothesized to affect brain health are drawn in part from Livingston et al. (20), updated to add brain stimulation via neurohumoral effects as a mechanism (15). This organization simplifies and facilitates psychoeducational efforts and helps older adults understand how to make evidence-informed choices about lifestyle changes. Education on how to evaluate medical information, such as the interpretation of clinical vs statistical significance and common risks indicators, such as odds ratios, gives older adults the tools they need to understand research findings and make informed choices. Finally, synthesizing the shared decision making and cognitive behavioral models is apt given both approach’s emphasis on psychoeducation and collaboration in developing goals and pursuing behavior change.

### Study design

All study procedures were completed remotely, by telephone, video conferencing or online data collection. Persons interested in the study contacted the investigators either by telephone or e-mail and were then screened by telephone using a standard script that assessed whether they met inclusion criteria. Those who were deemed potentially eligible were then asked to complete an assessment of their ability to participate in the online intervention by asking them to receive and open an e-mail with a link to a video conference and to open a link to an online questionnaire from the REDCap data capture software (21).

Participants who met these criteria were sent the informed consent document for review. In a second video conference, one of the investigators conducted the informed consent interview. Participants who chose to be in the study were asked to affirmatively indicate their consent for participation by opening another online questionnaire, checking a box on a form indicating their willingness to participate, and by providing an electronic signature. They were then asked to complete a baseline assessment comprising self-report questionnaires delivered by REDCap and a cognitive assessment using the CogniFit® (New York: Cognifit, Ltd.) platform. They were then scheduled for the first intervention session. These procedures were completed under a protocol approved by the Institutional Review Board of Nova Southeastern University.

### Participants

Participants were recruited from a local lifelong learning program that provided programming online during the pandemic. Several persons were referred by other participants and from a local religious organization. Inclusion criteria included being 50 years of age or older, having an interest in improving their brain health, and a willingness to participate in the 12-week study. Participants were required to have an e-mail address at which they could receive study communications and links to video conferences, and were required to be able to use videoconferencing software (San Jose, CA: Zoom Media).

### Measures

Measures used in this study included a variety of self-report measures assessing risk factors for cognitive decline and dementia, current health and emotional status, and cognitive functioning. At the conclusion of the study, participants completed a questionnaire that evaluated their views of the intervention with respect to its usefulness and usability and an interview that allowed them to provide feedback about it.

Participants completed a daily questionnaire assessing daily brain activities, the Cogstim index described below, such as exercise, healthy eating, and cognitive activity during weeks 1, 6, and 12 of the study. Finally, they also completed a second follow-up interview six weeks after completing the study.

### Primary Outcome Measures

The Australian National University Alzheimer’s Disease Risk Index (22) is a measure developed to allow an evidence-based assessment of individuals’ risk for developing Alzheimer’s disease. It includes assessments of demographic factors such as age and gender as well as education, medical history and status, depression, and cognitive and social activity. It has been validated on several cohorts of older adults (23).

The Memory Self-Efficacy Scale (24) is a brief scale distilled from a longer scale designed to assess older persons’ self-report of memory difficulties. It correlates well with the original scale and was independently validated in relation to list and text recall tasks.

The Dementia Knowledge Assessment Scale (25) is a 25-item measure developed to assess knowledge of dementia. It has been subjected to psychometric evaluation and found to have several subscales (26). In this study, we used its seven item Risk and Health Promotion subscale to assess participants’ knowledge of risk factors for dementia and strategies for improving their brain health.

### Secondary Outcome Measures

In addition to the primary outcome measures, participants completed several other self-report measures, including a measure of mood, the Center for Epidemiological Studies--Depression scale or CES-D (27), the 10-item version of the Perceived Stress Scale, or PSS (28), and the Cognitive Functioning and General Life Satisfaction scales drawn from the Patient-Reported Outcomes Measurement Information System, or PROMIS (29).

### Cogstim Index

Given increasing interest in using apps to track health status and behaviors (30), as part of this pilot study we sought to test a simple questionnaire that could be used to monitor brain health activities on a daily basis. The availability of this sort of questionnaire would allow interested individuals to track their ongoing efforts to develop brain healthy lifestyles over time, potentially enhancing their motivation to change their lifestyle and maintain those changes (31-33).

We developed a brief set of questions to enable daily tracking of cognitive training, other mentally stimulating activities such as reading or working puzzles, physical activity, and diet. Questions on cognitive training, mentally stimulating and physical activity simply ask the person whether they completed any of those activities. If so, the questionnaire was programmed to ask how many minutes were devoted to the activity and for a rating of the activity’s intensity. Scores for each activity were calculated as minutes multiplied by intensity. The items proposed by Sofi (34) were used to assess level of adherence to the Mediterranean diet. They were used because of their brevity and strong evidence base. As the questionnaire was based on a previously reported model emphasizing cognitive activity, brain stimulation, and reducing inflammation referred to as the Cogstim model (15), this index is referred to here as the Cogstim Index.

### Technology Acceptance Model Questionnaire

To assess participants’ views of the intervention, after completing the study they were asked to complete a questionnaire based on the Technology Acceptance Model (35). This scale provided estimates of participants’ perceptions of the usefulness, ease of use of the intervention, and likelihood of using it again. We hypothesized that if their views were positive, their average ratings of the intervention would be significantly more positive than the center rating point of a seven-point Likert-type rating scale.

### Interviews

Semi-structured interviews were completed during the week after the 12th session and again six weeks later. Participants’ responses to specific questions, such as whether they enjoyed the study and found it beneficial, were recorded. Open ended questions included, for example, “What did you like about the study?” and “How could we make it better?” Participant responses were recorded verbatim and entered in a spreadsheet for tabulation.

### Cognitive Training

Participants were given the opportunity to complete unlimited cognitive training using the commercial CogniFit® online software. This software was chosen because of substantial evidence showing that it is effective in improving cognitive function in older adults (36, 37) and because it could be used online by participants rather than in person. CogniFit® was also used for initial and follow-up assessments of cognitive functioning in several domains; the program provides an individually tailored training program based on participants’ initial assessments and ongoing progress. The program targets various cognitive functions, including attention, working memory, verbal and visual memory, and eye-hand coordination.

### Procedure

After obtaining informed consent, participants were e-mailed links to a series of online questionnaires and the cognitive assessment with the request that they complete both prior to the next scheduled visit. After completion of the baseline assessments, participants were scheduled for the first weekly session, an introductory overview of the program and goal setting. During this session, the Treatment as Usual (TAU) group received a general goal setting worksheet and were encouraged to choose goals to work on during the study. They were not otherwise encouraged to set specific goals in order to simulate what we hypothesize is a typical situation in which older persons are provided with information and encouraged to change their behavior without specific shared decision-making guidance or behavior change support.

Both groups were oriented to the CogniFit® software program and advised of its availability for use. This was to provide an estimate of the extent to which older adults, given a choice, would engage in computer-based cognitive training.

The CBSDM group received an overview of the model and were encouraged to choose activities from one of the three hypothesized brain health promoting mechanisms. They also received an introduction to SMART goal setting (38) and a structured goal setting worksheet. They were asked to develop at least one goal prior to the next weekly session.

In subsequent sessions, the investigator presented a 10-15-minute PowerPoint presentation on one of several topics, including risk and protective factors, meditation, sleep, stress management, exercise, cognitive training, Mediterranean diet, sense of purpose, social engagement, and mentally stimulating activities. Presentations usually included presentation of a relevant research study followed by suggestions for behavior change based on the study’s findings. For both groups, the final session was a review of all presentation topics and discussion of follow-up strategies.

For the TAU group, each session began with a general orientation to the topic, proceeded to the presentation, and concluded with answering questions and any discussion raised by the participants. For the CBSDM group, each session began with an agenda-setting slide in which the investigator proposed to review goals and progress and then to the topic presentation. The session concluded with a review of goals and, if needed, problem solving about progress toward goals.

As originally envisioned, the intervention was to have been delivered in small groups, but during the study it became clear that group scheduling would delay completing the pilot study unacceptably. Throughout the study the intervention was delivered one-on-one with one of the investigators (RLO).

After the 12 weekly sessions, participants again completed self-report measures and the cognitive assessment. They completed an exit interview based on a semi-structured format eliciting their views of the intervention, what they liked or disliked about it, and any suggestions for improvement. They were contacted again six weeks later again to be asked these questions and about the extent to which they had achieved their goals and maintained gains.

## Data Analyses

Data from self-report questionnaires were downloaded from REDCap and imported into the SPSS statistical package (IBM, Inc., Armonk, NY) for initial processing. Cognitive testing data were downloaded from the Cognifit website and imported into SPSS. Analyses were completed in several steps depending on the type of data. Mixed effects models were computed in R version 4.2.1 using the *lme4* package (39). Significance of treatment effects was assessed using the likelihood ratio test (40) comparing models with and without specific effects, including treatment group, time, and the interaction of the two. Effect sizes from likelihood ratio tests were converted to the more widely used *d* statistic using the R package *esc* (41). Given the observed variability of participants’ time completing cognitive training, in unplanned post-hoc analyses the effect of training more or less than one hour was also assessed in mixed effect models.

To assess ratings on the Technology Assessment Model subscales for Usefulness, Ease of Use, and Intent to Use Again, single sample *t*-tests were used to test the hypothesis that participant ratings would be significantly more positive than the scale midpoint of 3 on the 0 to 6 rating scales.

Ratings for each item in the Cogstim index completed in weeks 1, 6, and 12 were aggregated to provide mean values for each group and day. These values were summed to provide an overall index of daily brain health activities. Given the time between each set of one week ratings, self-reported brain health activities were evaluated using interrupted time series analysis in the Stata statistical software (Stata Corp, Inc., College Station, TX) using the *itsa* routine (42). Significance of observed values over time were assessed using Newey-West standard errors to account for autocorrelation with measurements over time.

For interviews, responses to questions with yes/no options were counted to allow tabulation. Responses to open ended questions were reviewed for the occurrence to similar responses to identify commonalities among them.

## Results

Participants included 18 persons 50 years of age and older of whom 16 completed the study. This group included 2 Black individuals and 16 White persons, with 6 men and 12 women, and three persons who stated their ethnic background was Hispanic. Participants’ interventions and data recording began in May 2021 and ended in October 2021. Descriptive data for participants in each treatment group are presented in Table 1, and a CONSORT diagram of participant flow is presented in Figure 1.

**Table 1.** Baseline Characteristics of Participants.

|  | TAU |  | CBSDM |  | TOTAL |  |  |  |  |
| --- | --- | --- | --- | --- | --- | --- | --- | --- | --- |
| Variable | Mean | SD | Mean | SD | Mean | SD | *t* | *p* | *d* |
| Education (years) | 18.50 | 1.51 | 17.40 | 2.32 | 17.89 | 2.03 | 1.16 | 0.27 | 0.55 |
| Age (years) | 72.00 | 5.53 | 73.20 | 9.55 | 72.67 | 7.83 | 0.31 | 0.76 | -0.15 |
| ADRI | 3.71 | 12.88 | 0.56 | 6.19 | 1.94 | 9.45 | 0.65 | 0.53 | 0.33 |
| DKAS | 5.13 | 1.13 | 4.60 | 1.35 | 4.83 | 1.25 | 0.88 | 0.39 | 0.42 |
| MSE | 47.63 | 8.98 | 54.10 | 5.59 | 51.22 | 7.79 | 1.88 | 0.08 | -0.89 |
| CESD | 15.75 | 9.87 | 13.10 | 8.82 | 14.28 | 9.12 | 0.60 | 0.56 | 0.29 |
| PSS | 11.75 | 8.45 | 12.40 | 6.60 | 12.11 | 7.25 | 0.18 | 0.86 | -0.09 |
| Life Satisfaction | 24.13 | 6.40 | 23.40 | 5.87 | 23.72 | 5.94 | 0.25 | 0.81 | 0.12 |
| Cognitive Function | 28.25 | 10.47 | 35.00 | 3.89 | 32.00 | 8.07 | 1.89 | 0.08 | -0.90 |

TAU = Treatment as Usual; CBSDM = Cognitive Behavioral Shared Decision Making; ADRI = Alzheimer's Disease Risk Inventory; DKAS = Dementia Knowledge Scale, Risk subscale; MSE = Memory Self-Efficacy Scale; CESD = Center for Epidemiological Studies Depression scale; PSS = Perceived Stress Scale; Life Satisfaction = PROMIS General Life Satisfaction Scale; Cognitive Function = PROMIS Cognitive Function Scale

**Figure 1.**
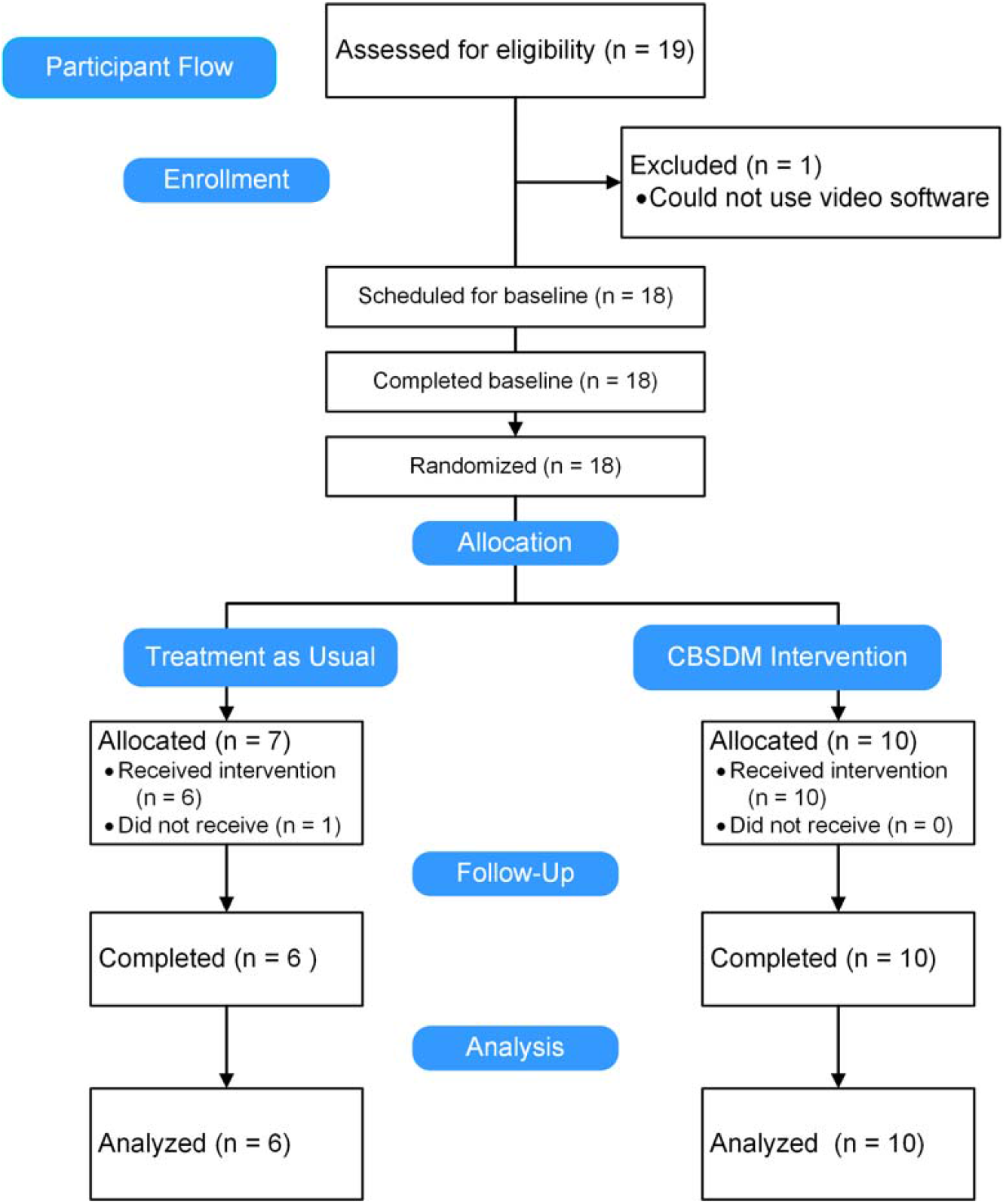
CONSORT Figure Showing Participant Flow.

## Primary Outcome Measures

No effects for the model with the Alzheimer’s Disease Risk Inventory were statistically significant, but inspection of the interaction plot suggested an increase in risk in the TAU group with no change in the CBSDM group (χ^2^ [1] = 0.30, *p* = 0.59, *d* = 0.27 [95th CI -0.71-1.26]) associated with a small effect size. For the Memory Self-Efficacy Scale, the effects of time, treatment group, and their interaction were not significant (all *p*s > 0.10).

For the Dementia Knowledge Assessment Scale, none of the effects were significant, but inspection of the interaction plot suggested a greater improvement in knowledge of risk factors in the CBSDM group (Figure 2). The effect of the interaction of treatment group by time from the likelihood ratio test (χ^2^ [1] = 1.19, *p* = 0.28, *d* = 0.63 [95th CI: -0.51-1.77) represented a moderate effect size.

**Figure 2.**
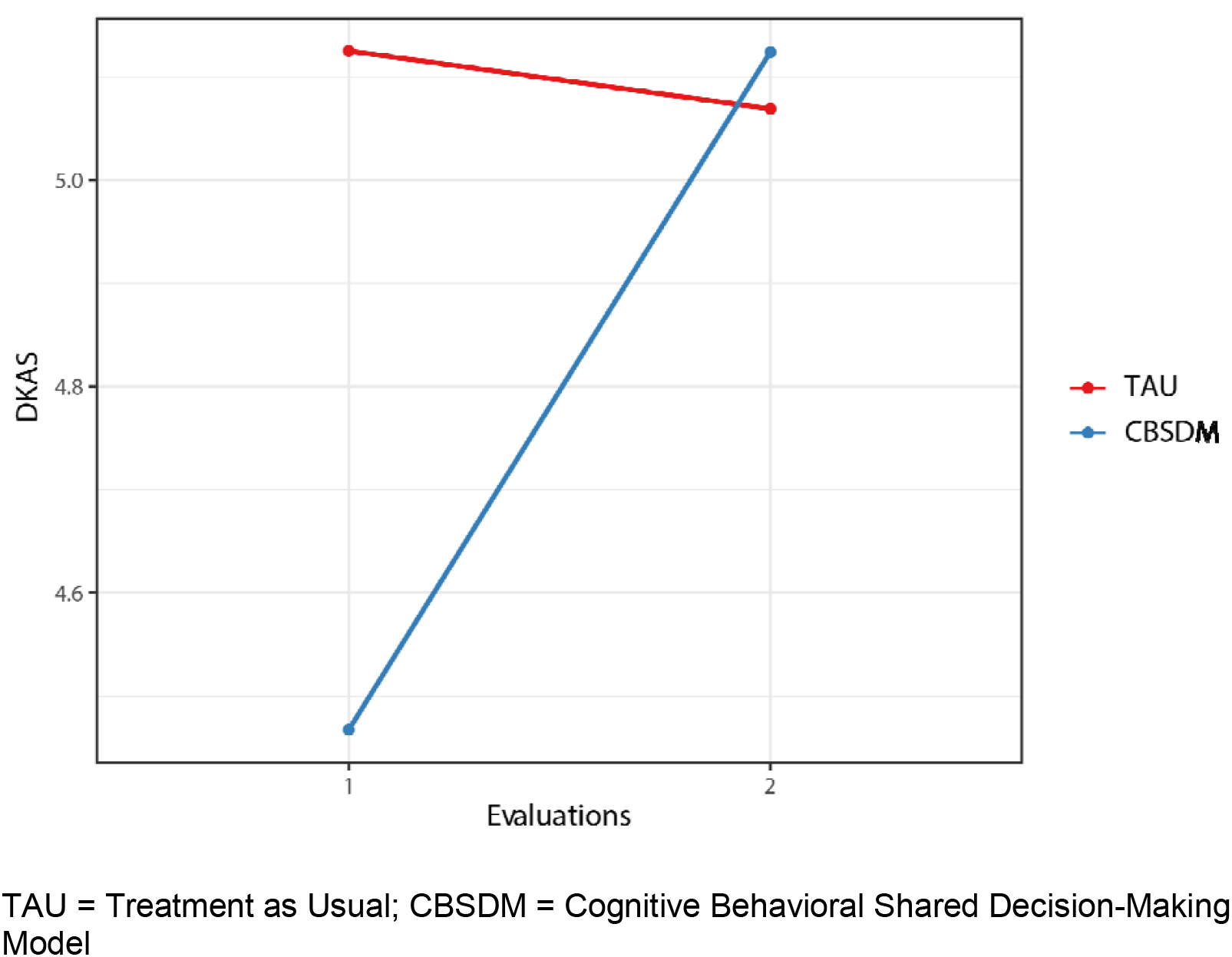
Dementia Knowledge Assessment Scale, Risk Subscale. TAU = Treatment as Usual; CBSDM = Cognitive Behavioral Shared Decision-Making Model

### Secondary Outcome Measures

For the PROMIS Life Satisfaction scale, effects for treatment, and the interaction of time with treatment group were not significant. The effect for time, however, approached conventional levels of significance and represented a large effect size (Figure 3; χ^2^ [1] = 3.37, *p* = 0.07, *d* = 1.03 [95th CI -0.07-2.14]). For the PROMIS self-report of Cognitive Function scale, no effects were significant. Inspection of the interaction plot suggested improvement in both groups during the study that represented a large effect size (χ^2^ [1] = 1.91, *p* = 0.16, *d =* 0.83 [95th CI -0.35-2.01].

**Figure 3.**
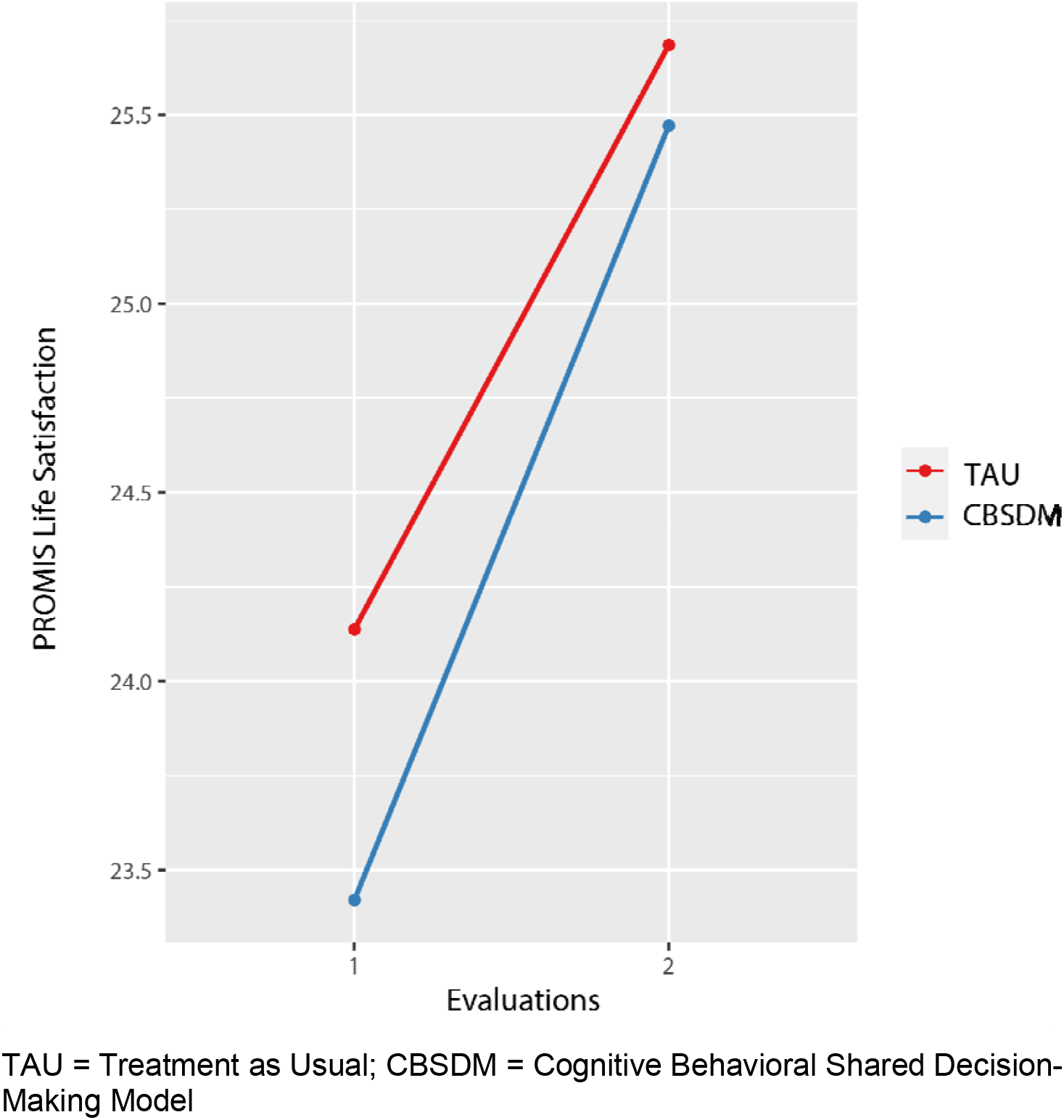
PROMIS Life Satisfaction Scale. TAU = Treatment as Usual; CBSDM = Cognitive Behavioral Shared Decision-Making Model

For the Center for Epidemiological Studies – Depression Scale, effects for treatment group and the interaction of group with time were not significant, but the decline in depressive symptoms during the study approached significance and represented a large effect size (χ^2^ [1] = 3.39, *p* = 0.07, *d* = 1.19 [95th CI -0.08-2.45]. For the Perceived Stress Scale, no model effects were statistically significant, although inspection of the interaction plot suggested a decrease in both groups that represented a medium effect size (χ^2^ [1] = 0.79, *p* = 0.37, *d* = 0.46 [95th CI -0.55-1.46].

### Cogstim Index

ITSA analyses for the overall questionnaire (summing scores for all domains) suggested increases in brain healthy activities for both groups in the second half of the study (weeks 6-12; *t* = 3.12, *p* = 0.004 for the CBSDM group and *t* = 2.73, *p* = 0.01 for the TAU group). For the physical activity score (Figure 4), the CBSDM group showed significant increases in the second half of the study (*t* = 2.44, *p* = 0.02) but the TAU group did not (*t =* 0.89, *p* = 0.38). The difference between the two groups, however, was not significant (*t* = 0.40, *p* = 0.69).

**Figure 4.**
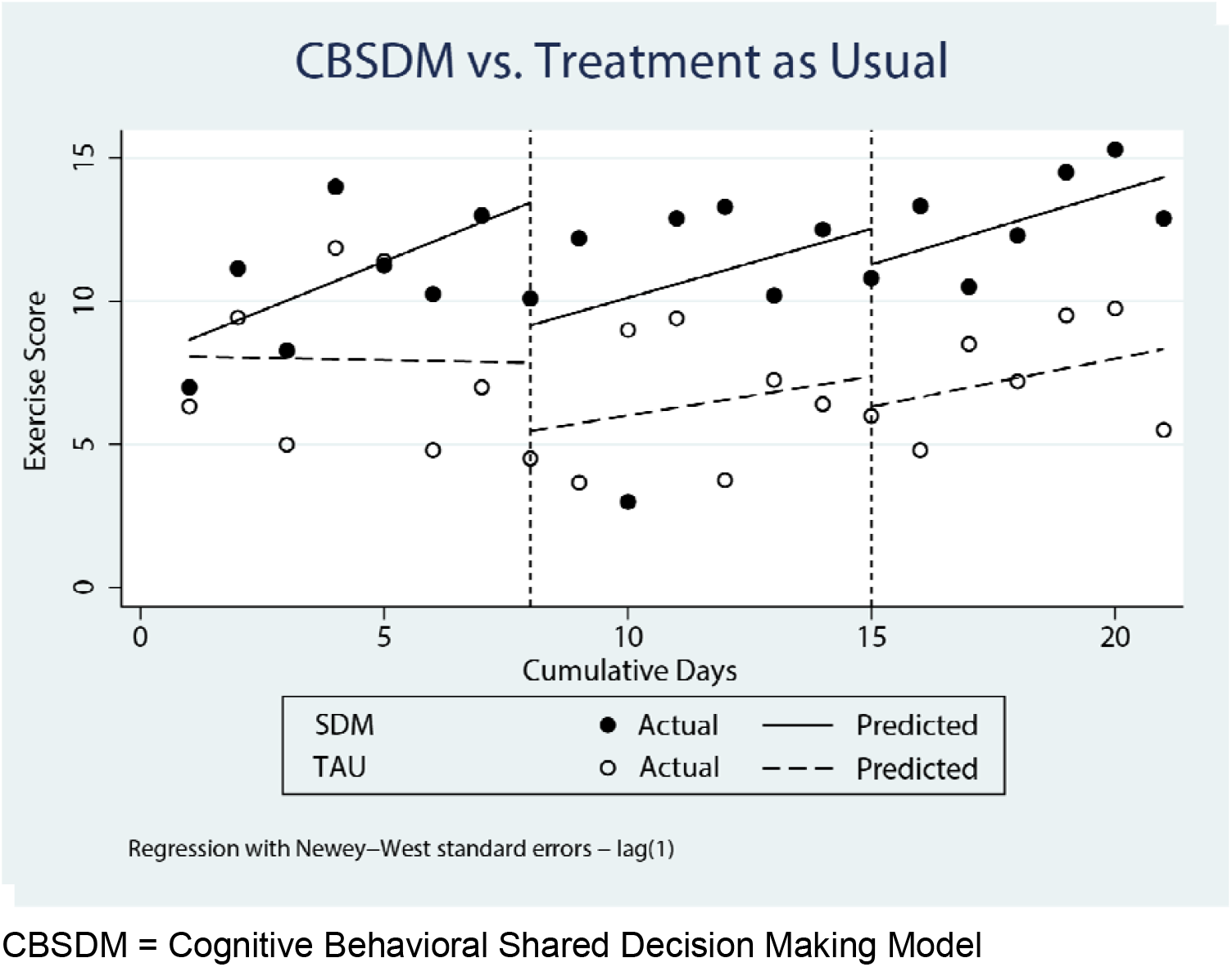
Interrupted Time Series Analysis for Physical Activity in Weeks 1, 6, and 12. CBSDM = Cognitive Behavioral Shared Decision Making Model

### Cognitive Assessments

Evaluation of training effects on the Cognifit® cognitive measures before and after the study showed, in general, evidence of modest but nonsignificant improvements in cognitive domain scores. When time spent training was included in models, however, the effect of time approached significance and represented a large effect size (e.g., for attention, χ^2^[1] = 2.88, *p* = 0.09, *d* = 0.93 [95th CI -0.14-2.02]; for working memory, χ^2^ [1] = 2.46, *p* = 0.12, *d* = 1.07 [95th CI -0.04-2.19]). This finding suggests that time spent in cognitive training had a positive impact on subsequent performance on cognitive measures.

Results of unplanned comparisons of those who completed at least one hour of cognitive training compared to those who did not were consistent with this interpretation of the impact of training on posttest scores, with a significant time by group effect (trained vs. not) on the motor coordination subtest that was associated with a very large effect size (χ^2^ [1] = 9.57, *p =* 0.002, *d* = 2.44 [95th CI 0.89-3.98]). This finding supports the efficacy of training on this skill for those who engaged in one hour or more of training.

### Acceptability and Usability

Participants’ ratings of the intervention on all TAM subscales of the Technology Acceptance Model scale were significantly different from the scale’s midpoints, with ratings of Usefulness, Ease of Use, and Intent to Use subscales in the future representing very large effect sizes (*d =* 3.50 [95th CI 1.94-5.04], 2.56 [95th CI 1.35-3.74], and 1.69 [95th CI 0.78-2.58, respectively). No between-group differences were significant.

### Interviews

In interviews completed during the week after the intervention, all participants stated that they found the program helpful (n = 16) and that they enjoyed it (n = 16). All but one indicated that they would do it again. One participant said, “the program was extremely valuable,” while another stated, “I was made keenly aware of the need to keep my mind active.” Several indicated that they found weekly sessions helpful, especially the structure of linking discussions of topical presentations with specific research studies ensuring that there was an empirical foundation for lifestyle recommendations.

Although many of the participants indicated that they were aware of the information presented in the weekly sessions, they noted that hearing the topics again reinforced for them the importance of a brain healthy lifestyle and helped them maintain a focus on making lifestyle changes, as in “taking concrete steps” to develop better health habits. Finally, several of the participants in the CBSDM model group noted that the structure of the sessions, with goal setting and follow-up was helpful to them in initiating behavior changes.

In interviews six weeks after intervention completion, participants continued to state that they had found the program helpful and that they enjoyed it (16 of 16). When asked if they had continued to work on their goals, 15 of 16 stated that they were continuing to work on changing their behavior. Four participants indicated that they had achieved their brain health goals, two said they had not, and 10 said they had partially achieved their goals. Fourteen of 16 stated they would do the program again.

## Discussion

The purpose of this study was to assess the acceptability, usability, and efficacy of a cognitive behavioral shared decision-making model-based intervention in helping older adults develop more brain healthy lifestyles. The model was developed in order to help older adults make evidence-informed choices about activities that might benefit them and to give them the tools they required to initiate and sustain behavior change. Results of the study showed that the content and format of the intervention were acceptable and useful to the participants, and that the content and format of the CBSDM based intervention supported efforts at behavior change. Results were consistent with greater improvement in knowledge of dementia risk factors in participants in the CBSDM group, as well as suggesting greater change in behavior over time in that group. It may also be noted that participants in both groups appear to at least subjectively have benefited from participation, with both groups showing substantial decreases in stress and depressive symptoms while showing increases in life satisfaction and self-report of cognitive function.

The online format of the intervention as well as all study procedures was welcome to many of our participants during a time when there were high levels of concern about face-to-face contact due to the COVID epidemic. The study showed that even for participants who were older than 65, the online format was acceptable. The online format also provided a useful degree of flexibility to several participants who traveled during the study and were able to attend sessions even when they were not at home.

Several limitations of the study should be acknowledged. While our highly-educated sample of participants were able to use the technology underlying online interactions, including regular video sessions as well as routine communication by e-mail, it is not clear that all other older adults would be able to do so. A second important limitation of the study was the format of the individual coaching sessions, as all intervention sessions were delivered by a single person (RLO). Although the structure of each session for both intervention groups was established by following previously prepared PowerPoint slide sets, no formal monitoring strategy was employed to assess intervention fidelity. A second drawback of the individual session format was its time-consuming nature; the person conducting the sessions provided over 80 hours of direct content delivery to the participants. While several participants expressed appreciation at receiving this level of attention, it would be difficult if not impossible to deploy the intervention more widely. In the future, we plan to explore ways to automate some content delivery by way of an online course and to provide group online sessions to support participants’ behavior change efforts.

In summary, results of this study show that the CBSDM model-based intervention was positively viewed by older adults who found its content and format helpful to them in developing more brain healthy lifestyles. While limited by the small sample size, findings of improvements in knowledge and changes in behavior favoring the CBSDM group as well as participants’ comments suggest that the structured format in this group with its emphasis on goal setting and self-monitoring may be especially useful in helping older adults change their behavior.

## Data Availability

All data produced in the present study are available upon reasonable request to the authors

## Acknowledgments

The authors declare no acknowledgements.

## Author contributions

Raymond L. Ownby, designed the research, conducted the research, analyzed the data and wrote the paper. He had primary responsibility for the final content of the manuscript. Rosemary Davenport, ARNP, assisted in study design and conduct of research and helped edit the paper. Both authors read and approved the final manuscript.

## References

1. van Dyck CH, Swanson CJ, Aisen P, Bateman RJ, Chen C, Gee M, et al. Lecanemab in early Alzheimer’s disease. New England Journal of Medicine. 2022;388(1):9–21. DOI: 10.1056/NEJMoa2212948

2. Zissimopoulos J, Crimmins E, St Clair P. The value of delaying Alzheimer’s disease onset. Forum Health Econ Policy. 2014;18(1):25–39. DOI: 10.1515/fhep-2014-0013

3. Lock SL. The benefits of brain health to our economies. Nature Aging. 2023;3(1):1–2. DOI: 10.1038/s43587-022-00302-z

4. Livingston G, Huntley J, Sommerlad A, Ames D, Ballard C, Banerjee S, et al. Dementia prevention, intervention, and care: 2020 report of the Lancet Commission. The Lancet. 2020;396(10248):413–46. DOI: 10.1016/S0140-6736(20)30367-6

5. Ngandu T, Lehtisalo J, Solomon A, Levälahti E, Ahtiluoto S, Antikainen R, et al. A 2 year multidomain intervention of diet, exercise, cognitive training, and vascular risk monitoring versus control to prevent cognitive decline in at-risk elderly people (FINGER): a randomised controlled trial. The Lancet. 2015;385(9984):2255–63. DOI: 10.1016/S0140-6736(15)60461-5

6. Hellmuth J. Can we trust The End of Alzheimer’s? The Lancet Neurology. 2020;19(5):389–90. DOI: 10.1016/S1474-4422(20)30113-7

7. Mehegan L, Rainville C. 2021 AARP aurvey on the perceptions related to a dementia diagnosis: Adults age 40+. . Washington DC: AARP Research; 2021.

8. Zülke AE, Luppa M, Köhler S, Riedel-Heller SG. Knowledge of risk and protective factors for dementia in older German adults A population-based survey on risk and protective factors for dementia and internet-based brain health interventions. PLOS ONE. 2022;17(11):e0277037. DOI: 10.1371/journal.pone.0277037

9. Friedman BB, Suri S, Solé-Padullés C, Düzel S, Drevon CA, Baaré WFC, et al. Are people ready for personalized brain health? Perspectives of research participants in the Lifebrain Consortium. Gerontologist. 2020;60(6):1050–9. DOI: 10.1093/geront/gnz155

10. Friedman DB, Laditka JN, Hunter R, Ivey SL, Wu B, Laditka SB, et al. Getting the message out about cognitive health: A cross-cultural comparison of older adults’ media awareness and communication needs on how to maintain a healthy brain. The Gerontologist. 2009;49(S1):S50–S60. DOI: 10.1093/geront/gnp080

11. Wilcox S, Sharkey JR, Mathews AE, Laditka JN, Laditka SB, Logsdon RG, et al. Perceptions and beliefs about the role of physical activity and nutrition on brain health in older adults. The Gerontologist. 2009;49(S1):S61–S71. DOI: 10.1093/geront/gnp078

12. Maust DT, Solway E, Langa KM, Kullgren JT, Kirch M, Singer DC, et al. Perception of dementia risk and preventive actions among US adults aged 50 to 64 years. JAMA Neurology. 2020;77(2):259–62. DOI: 10.1001/jamaneurol.2019.3946

13. Elwyn G, Frosch D, Thomson R, Joseph-Williams N, Lloyd A, Kinnersley P, et al. Shared decision making: A model for clinical practice. J Gen Intern Med. 2012;27(10):1361–7. DOI: 10.1007/s11606-012-2077-6

14. Hibbard JH. Engaging health care consumers to improve the quality of care. Medical Care. 2003;41(1):I-61-I-70, https://journals.lww.com/lww-medicalcare/Fulltext/2003/01001/Engaging_Health_Care_Consumers_to_Improve_the.7.aspx

15. Ownby RL, Waldrop D. Cogstim: A shared decision-making model to support older adults’ brain health. Current Alzheimer Research. 2023;20(Epub ahead of print). DOI: 10.2174/1567205020666230525110814

16. Bredesen DE, Amos EC, Canick J, Ackerley M, Raji C, Fiala M, et al. Reversal of cognitive decline in Alzheimer’s disease. Aging. 2016;8(6):1250–8. DOI: 10.18632/aging.100981

17. Small G, Vorgan G. The Alzheimer’s prevention program. New York: Workman; 2012.

18. Sherzai D, Sherzai A. The Alzheimer’s solution. London: Simon & Schuster UK; 2017.

19. Beck JS. Cognitive behavior therapy: Basics and beyond (3rd ed.). New York: Guilford; 2020.

20. Livingston G, Sommerlad A, Orgeta V, Costafreda SG, Huntley J, Ames D, et al. Dementia prevention, intervention, and care. The Lancet. 2017;390(10113):2673–734. DOI: 10.1016/S0140-6736(17)31363-6

21. Harris PA, Taylor R, Thielke R, Payne J, Gonzalez N, Conde JG. Research electronic data capture (REDCap)--a metadata-driven methodology and workflow process for providing translational research informatics support. J Biomed Inform. 2009;42(2):377–81. DOI: S1532-0464(08)00122-6 [pii];10.1016/j.jbi.2008.08.010 [doi]

22. Anstey KJ, Cherbuin N, Herath PM. Development of a new method for assessing global risk of Alzheimer’s disease for use in population health approaches to prevention. Prev Sci. 2013;14(4):411–21. DOI: 10.1007/s11121-012-0313-2

23. Anstey KJ, Cherbuin N, Herath PM, Qiu C, Kuller LH, Lopez OL, et al. A self-report risk index to predict cccurrence of dementia in three independent cohorts of older adults: The ANU-ADRI. PLOS ONE. 2014;9(1):e86141. DOI: 10.1371/journal.pone.0086141

24. Zelinski EM, Gilewski MJ. A 10-item Rasch modeled memory self-efficacy scale. Aging Ment Health. 2004;8(4):293–306. DOI: 10.1080/13607860410001709665

25. Annear MJ, Toye CM, Eccleston CE, McInerney FJ, Elliott K-EJ, Tranter BK, et al. Dementia Knowledge Assessment Scale: Development and preliminary psychometric properties. Journal of the American Geriatrics Society. 2015;63(11):2375–81. DOI: 10.1111/jgs.13707

26. Annear MJ, Toye C, Elliott KJ, McInerney F, Eccleston C, Robinson A. Dementia Knowledge Assessment Scale (DKAS): Confirmatory factor analysis and comparative subscale scores among an international cohort. BMC Geriatr. 2017;17(1):168. DOI: 10.1186/s12877-017-0552-y

27. Radloff LS. The CES-D Scale: A self-report depression scale for research in the general population. Applied Psychological Measurement. 1977;1:385–401,

28. Cohen S, Janicki-Deverts D. Who’s stressed? Distributions of psychological stress in the United States in probability samples from 1983, 2006, and 2009. Journal of Applied Social Psychology. 2012;42(6):1320–34. DOI: doi: 10.1111/j.1559-1816.2012.00900.x

29. Cella D, Riley W, Stone A, Rothrock N, Reeve B, Yount S, et al. The Patient-Reported Outcomes Measurement Information System (PROMIS) developed and tested its first wave of adult self-reported health outcome item banks: 2005-2008. J Clin Epidemiol. 2010;63(11):1179–94. DOI: 10.1016/j.jclinepi.2010.04.011

30. McKay FH, Wright A, Shill J, Stephens H, Uccellini M. Using health and well-being apps for behavior change: A systematic search and rating of apps. JMIR Mhealth Uhealth. 2019;7(7):e11926. DOI: 10.2196/11926

31. Harkin B, Webb TL, Chang BP, Prestwich A, Conner M, Kellar I, et al. Does monitoring goal progress promote goal attainment? A meta-analysis of the experimental evidence. Psychol Bull. 2016;142(2):198–229. DOI: 10.1037/bul0000025

32. Dunn EE, Gainforth HL, Robertson-Wilson JE. Behavior change techniques in mobile applications for sedentary behavior. Digit Health. 2018;4:2055207618785798. DOI: 10.1177/2055207618785798

33. Compernolle S, DeSmet A, Poppe L, Crombez G, De Bourdeaudhuij I, Cardon G, et al. Effectiveness of interventions using self-monitoring to reduce sedentary behavior in adults: A systematic review and meta-analysis. International Journal of Behavioral Nutrition and Physical Activity. 2019;16(1):63. DOI: 10.1186/s12966-019-0824-3

34. Sofi F, Macchi C, Abbate R, Gensini GF, Casini A. Mediterranean diet and health status: An updated meta-analysis and a proposal for a literature-based adherence score. Public Health Nutr. 2014;17(12):2769–82. DOI: 10.1017/s1368980013003169

35. Venkatesh V. Determinants of perceived ease of use: Integrating control, intrinsic motivation, and emotion into the Technology Acceptance Model. Information Systems Research. 2000;11(4):342,

36. Shah TM, Weinborn M, Verdile G, Sohrabi HR, Martins RN. Enhancing cognitive functioning in healthly older adults: A systematic review of the clinical significance of commercially available computerized cognitive training in preventing cognitive decline. Neuropsychology Review. 2017;27(1):62–80. DOI: 10.1007/s11065-016-9338-9

37. Shatil E. Does combined cognitive training and physical activity training enhance cognitive abilities more than either alone? A four-condition randomized controlled trial among healthy older adults. Front Aging Neurosci. 2013;5:8. DOI: 10.3389/fnagi.2013.00008

38. Doran GT. There’s a S.M.A.R.T. way to write managements’s goals and objectives. Management Review. 1981;70(11):35–6,

39. Bates D, Maechler M, Bolker B, Walker S. Fitting linear mixed-effects models using lme4. Journal of Statistical Software. 2015;67(1):1–48. DOI: doi:10.18637/jss.v067.i01.

40. Luke SG. Evaluating significance in linear mixed-effects models in R. Behav Res Methods. 2017;49(4):1494–502. DOI: 10.3758/s13428-016-0809-y

41. Ludecke D. Effect size computation for meta analysis. Vienna, Austria: R Core Team; 2019 DOI: 10.5281/zenodo.1249218

42. Linden A. ITSA: Stata module to perform interrupted time series analysis for single and multiple groups. Boston, MA: Boston College Department of Economics; 2014 DOI, https://ideas.repec.org/c/boc/bocode/s457793.html

